# Three year experience of Upper Gastrointestinal Endoscopic procedures at a tertiary care hospital of South Punjab

**DOI:** 10.1101/2021.11.11.21266215

**Authors:** Farooq Mohyud Din Chaudhary, Muhammad Asif Gul, Rizwan Hameed, Muhammad Ilyas, Muhammad Zubair, Qasim Umar, Syeda Manal Altaf, Ahsan Tameez-ud-din

## Abstract

**Introduction:** Endoscopy has become a necessity in diagnosing gastrointestinal (GI) disorders. The objective of our study was to evaluate the different indications and findings of upper GI endoscopy.

**Methods:** This retrospective analysis was undertaken at department of Gastroenterology, Nishtar Hospital Multan. Records of all upper GI endoscopic procedures from 1st January 2018 till 31st December 2020 were evaluated.

**Results:** A total 3299 upper GI endoscopic procedures were perfumed during the three-year time period. Mean age was 47 years. Majority of patients were males. Almost 48% of patients belonged to the middle-aged group. The most common indication was upper GI bleeding (57%), followed by dyspepsia (15%). The most common finding was esophageal varices (43%), followed by portal gastropathy (26%) and gastritis (16%).

**Conclusion:** This study concludes that the majority of endoscopies are being undertaken as a result of complications of cirrhosis and portal hypertension.

## INTRODUCTION

The first fibreoptic gastroscopy was introduced by Larry Curtiss in 1958 [1]. That was the start of a new era in diagnostic evaluation of gastrointestinal (GI) problems. Since then, Barium studies have been almost completely replaced by Endoscopy [2]. Endoscopy is the best modality to evaluate upper gastrointestinal (GI) mucosa. Rapid technological advances have led to remarkable changes in diagnosis and management of GI problems. Endoscopy is superior to any modality as it allows direct visualization of the mucosal surfaces of the esophagus, stomach and proximal duodenum [3].

Abdominal problems including upper GI disorders have high morbidity and mortality in the range of 2% to 33% [4, 5]. Upper GI endoscopy has both diagnostic and therapeutic potential for gastrointestinal disorders [6]. Among Upper GI problems, bleeding is a fatal medical emergency. Upper GI bleeding has a wide variety of causes all around the world including but not limited to peptic ulcers, esophageal varices, gastric erosions, and mucosal tear [7]. In a developing country like Pakistan, there is a high burden of chronic liver disease (CLD) due to infection with hepatitis B and C viruses. As a result of it, there is increased frequency of variceal bleeding in this region. Varices are seen in approximately 30% of compensated and 60% of decompensated CLD patients [8]. Endoscopic services are yet not readily accessible or affordable in most healthcare facilities [9]. Moreover, scanty data is available about demographics, indications, and findings of UGI endoscopy in Pakistan. We, therefore, aim to perform a study to document demographic characteristics, indications, and endoscopic findings in patients undergoing UGI endoscopy at a tertiary care hospital in Multan, which has a large number of referrals from around the region.

## METHODS

### Scope of study

This retrospective study was carried out at the Department of Gastroenterology, Nishtar Hospital Multan. This study was approved by the Institutional Review Board of Nishtar Medical University Multan (Reference No. 18674). To understand the spectrum of endoscopic activity, record of all EGDs performed from 1st January 2018 till 31st December 2020 were investigated.

### Data Collection

Using non-probability, consecutive sampling, all the patients who underwent endoscopic procedures during this time period were included in our study. From the endoscopy record register patient’s age, gender, type of procedure, indication of procedure and findings of endoscopy were noted. Confidentiality of patients was ensured.

### Data Analysis

Data were entered and evaluated in Statistical Package for Social Sciences (SPSS) version 20 (IBM Corp, Armonk, US). The results were reported as frequencies, percentages, and tables.

## RESULTS

A total of 3299 patients underwent EGD during our study time period. Out of the total patients enrolled in our study, there were 1941 males (58.8%) and 1358 females (41.2%). Mean age of study population was 47.15 years with a standard deviation of 15.98 years. The youngest patient who underwent EGD in our center was 10 years old, while the eldest one was 99 years old. Table 1 shows age wise distribution of study population. It can be seen that a majority (48%) of patients belonged to the middle age group of 41-60 years.

**Table 1:** Age wise distribution of patients undergoing EGD.

| **Age group**<br>**(years)** | **Frequency** | **Percent** |
| --- | --- | --- |
| ≤ 20 | 213 | 6.5 |
| 21-40 | 915 | 27.7 |
| 41-60 | 1581 | 47.9 |
| 61-80 | 551 | 16.7 |
| >80 | 39 | 1.2 |

Table 2 shows the different indications for which EGD was performed. As evident the most common indication was upper GI bleeding, accounting for more than half of our study population. Persistent dyspepsia was the second most common indication for EGD (15%). Table 3 shows the spectrum of endoscopic findings in patients who underwent EGD. Esophageal varices were the most common endoscopic findings seen in 43.6% of our study population. Portal hypertensive gastropathy was the second most common EGD finding accounting for 26%.

**Table 2:**
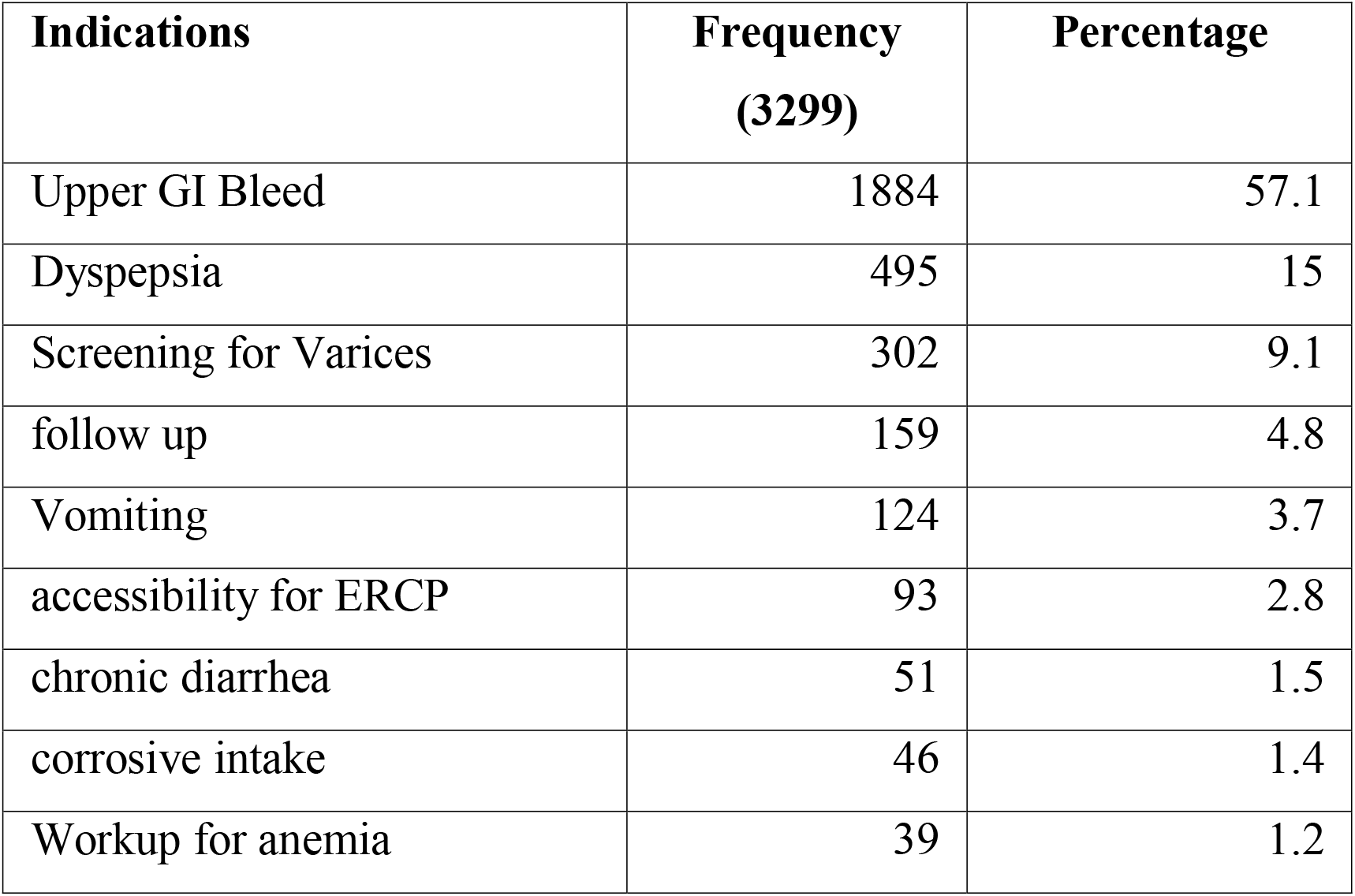
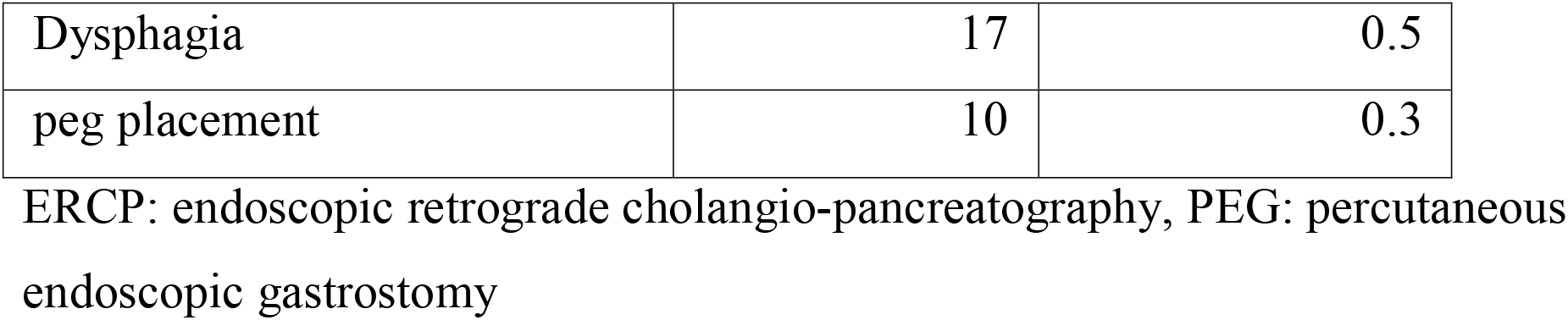
Different indications for EGD.

**Table 3:** Endoscopic findings.

| **EGD Findings** | **Frequency**<br>**(3299)** | **Percentage** |
| --- | --- | --- |
| Esophageal varices | 1439 | 43.6 |
| Portal gastropathy | 872 | 26.4 |
| Gastritis | 570 | 17.3 |
| Normal EGD | 334 | 10.2 |
| Incompetent LES | 163 | 4.9 |
| Fundal Varices | 91 | 2.8 |
| Hiatus Hernia | 66 | 2.0 |
| Gastric Growth | 61 | 1.85 |
| Esophageal growth | 58 | 1.76 |
| Esophageal Ulcer | 40 | 1.2 |
| Gastric Ulcer | 32 | 1.0 |
| Duodenal Ulcer | 30 | 0.9 |
| Esophageal candidiasis | 26 | 0.80 |
| Duodenitis | 26 | 0.80 |
| Pyloric Stricture | 24 | 0.72 |
| Loss of regular folds of stomach | 20 | 0.61 |
| Esophageal web | 15 | 0.45 |
| Reflux Esophagitis | 14 | 0.42 |
| Corrosive Injury | 12 | 0.36 |
| Esophageal Stricture | 11 | 0.33 |
| PEG placement | 10 | 0.30 |
| Ampullary mass | 8 | 0.24 |
| Duodenal growth | 4 | 0.12 |
| Gastric polyp | 3 | 0.10 |
| Celiac disease | 2 | 0.06 |
| Duodenal polyps | 2 | 0.06 |
| Worms | 2 | 0.06 |
| Duodenal stricture | 2 | 0.06 |
| Gastric polyps | 1 | 0.03 |
| Mallory Weis Tear | 1 | 0.03 |

LES: lower esophageal sphincter

## DISCUSSION

Upper GI endoscopy is of paramount importance in the diagnosis and management of gastrointestinal diseases. This was the first time an endoscopy audit was done at our hospital. There were significantly more male patients who underwent EGD as compared to females. This was in contrast to a local study in Rawalpindi [10] and other international studies [11] where female gender was more in number as compared to male population who underwent EGD. Another interesting observation was that frequency of normal EGD was found to be only 10.1 % in contrast to 61% in a local study and 24 % reported in a Canadian study[10, 12). Whether this difference is due to strict adherence to appropriate indications of procedure or reluctance of endoscopists in giving normal endoscopy report is not clear.

Upper GI bleed was the most common indication for EGD where 57% of procedures were done for this reason which is significantly high in comparison with international study where 14 % of upper GI endoscopy procedures had upper GI bleed as indication [12].The most common EGD finding was esophageal varices and portal gastropathy, present in 43 and 26 percent of patients, respectively. This is in contrast to a local study where esophageal varices were found in almost 16% of patients [10]. Gastritis was found in 17% of patients in our study, which is comparable to a local study (15%), however a lot less than an international study which had almost 38% gastritis [12]. Other less common causes include gastric ulcers, esophageal and gastric malignancy, duodenal ulcers and Mallory Weiss tear. The results are comparable to other local studies.

Dyspepsia was another common indication (15%) for carrying out upper GI endoscopies. However, it was a lot less as compared to an international study in which almost 34% patients underwent EGD due to dyspepsia [12].

## CONCLUSION

This three-year endoscopy audit confirms that the majority of EGDs are being performed for complications of cirrhosis and portal hypertension. Upper GI bleeding is the most common indication and esophageal varices are the most common finding on EGD in this part of the world. Efforts should be undertaken to eliminate the root cause of these problems like viral hepatitis B and C, so that liver decompensation can be prevented. Moreover the endoscopists should always be ready to anticipate variceal bleeding in this part of the world.

## Supporting information

IRB approval certificate

## Data Availability

All data produced in the present work are contained in the manuscript.

