## Supplementary figures and images for "Three year experience of Upper Gastrointestinal Endoscopic procedures at a tertiary care hospital of South Punjab"

### IRB approval certificate

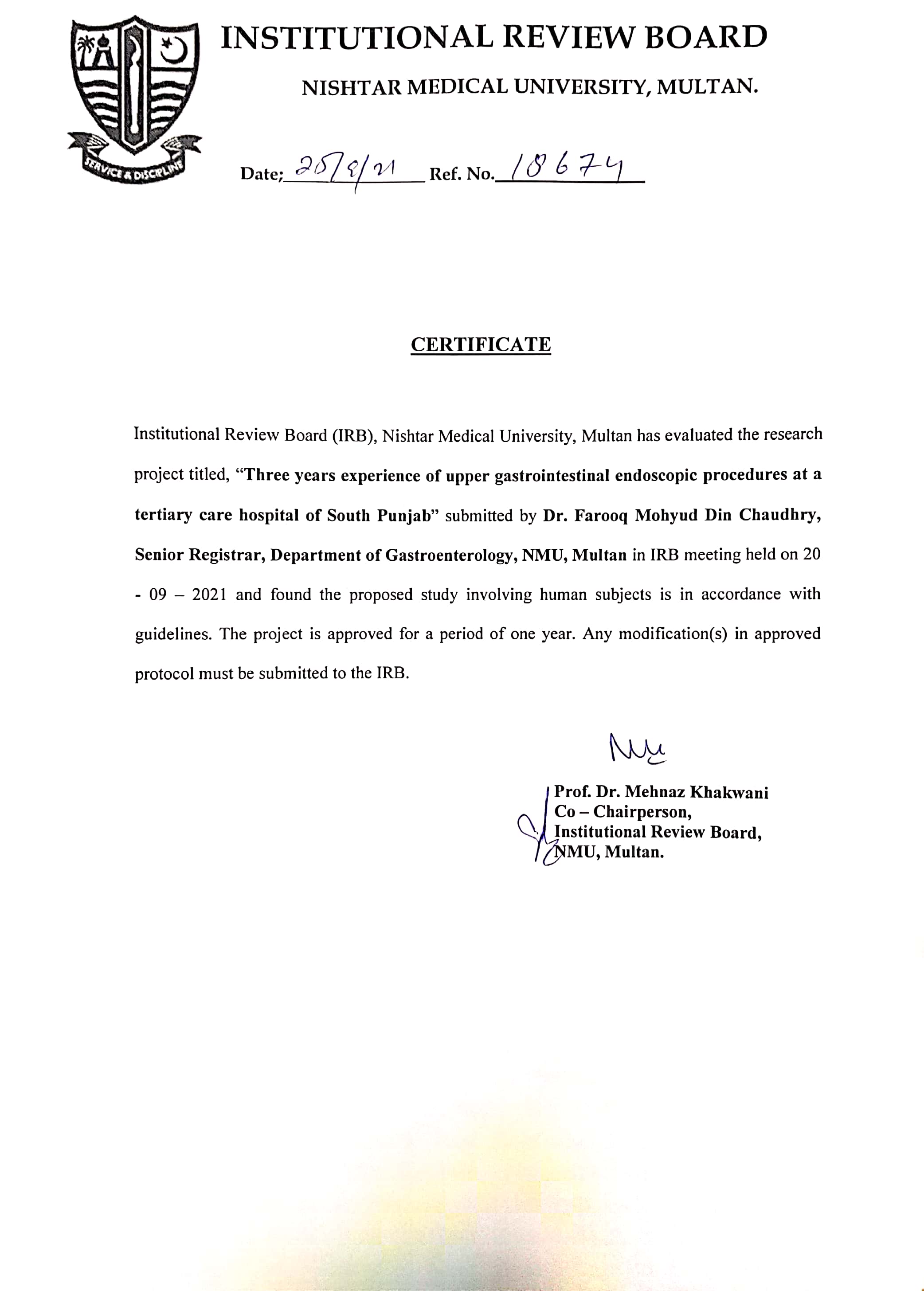
